# Association of Polymorphic Variants of HHIP, ADRB2 and IL-33 Genes with Clinical Manifestations of Bronchial Asthma in Children

**DOI:** 10.1101/2023.12.14.23299853

**Authors:** Yu.S. Alieva, E.G. Furman, E.I. Kondratyeva, E.V. Loshkova, V.S. Sheludko, V.S. Sokolovsky, M.S. Ponomareva, E.A. Khuzina, R.S. Aushova

## Abstract

Studying the contribution of genetic mechanisms to the development of bronchial asthma (BA) is to look for associations of the disease and its phenotypes with polymorphic markers of candidate genes.

**Objective:** To investigate the association of polymorphic variants of HHIP, ADRB2 and IL-33 genes with the phenotypes of clinical course of BA in children and the effectiveness of therapy of the disease

**Patients and methods:** A cohort single-center study of 90 bronchial asthma patients aged 5 to 17 years with an established diagnosis of bronchial asthma of varying degree of severity and control was conducted. The allele and genotype frequencies of polymorphic loci of the following genes were studied: rs12551256-A and rs146597587-G of IL-33 gene; rs12504628 of HHIP gene and ARG16GLY rs1042713 of ADRB2 gene in 90 BA patients with regard to the severity and control of the disease. In children with severe BA, as well as in children with poorly controlled/uncontrolled asthma (n=26), sequencing of the entire coding sequence of the IL-33 gene located on the 9th chromosome in the 9p24.1 region was additionally performed (search for mutations in 9 exons).

**Results:** Comparison of genetic markers in patients with severe BA (tBA) and non-severe BA (nBA) revealed a reduced risk of severe disease realization among those carrying the TT genotype (OR=0.221 (95% CI: 0.059-0.828; χ2=5.759; p=0.056)) and the T allele (OR=0.491 (95% CI: 0.190-1.269; χ2=4.270; p=0.039)) of the studied genetic variant rs12504628 (T>C) of the HHIP gene, the frequency of the CC genotype in severe BA was 64%, versus 28% in nonsevere BA, and the C allele 77% versus 52%.Comparison of genetic markers in patients with a combination of atopic dermatitis (AtD) and bronchial asthma (BA+AtD) and BA without AtD (BA without AtD) revealed an increased risk of combining asthma and dermatitis among individuals carrying the TT genotype (OR=2.875 (95% CI: 1.130-7.316; χ2=5.751; p=0.056)) of genetic variant rs12504628 (T>C) of the HHIP gene. Sequencing and exome analysis of the IL-33 gene showed a statistically significant positive association between the frequency of lesions in exons 4 (r=0.417; p=0.034) and 6 (r=0.593; p=0.001) on the one hand and the severity of BA on the other. Nucleotide substitutions in these exons were found to be more frequently associated with the severe course of bronchial asthma.

**Conclusion:** It was shown that TT genotype of genetic variant rs12504628 (T>C) of HHIP gene reduces the risk of severe BA, but increases the risk of atopic dermatitis combined with BA by 2.8 times. The CC+ST genotype of the HHIP gene increases the risk of drug allergy against the background of BA by 2.9 times. Polymorphic variants in exons 4 and 6 of the IL-33 gene are more often combined with moderate and severe asthma, and nucleotide substitutions in exons 4 and 6 are associated with a severe course of BA.

## Introduction

Bronchial asthma (BA) is one of the most important problems of modern theoretical and practical medicine. According to the report of The Global Asthma Network, currently, about 348 million people suffer from this disease, at least 14% of them are children [1]. It is estimated that in 2019, the number of patients with BA was 262 million and 461,000 deaths from the disease were reported [2].

BA is a heterogeneous disease based on chronic airway inflammation characterized by reversible bronchoobstructive syndrome, recurrent episodes of wheezing, dyspnoea, chest congestion and cough. The symptoms vary in time and intensity [3, 4]. The main goals of treatment of patients with BA are to achieve and maintain optimal control of the disease and to prevent exacerbations [5].

Despite a wide BA availability of inhaled glucocorticosteroids (iGCS) and standardized guidelines for asthma management, disease control remains suboptimal in most children. More than 50% of all children with BA experience at least one exacerbation each year, including children with non-severe asthma. In Russia, the problem of asthma control is also very urgent, as only 23% of patients achieve complete control of the disease. Among the reasons for insufficient control of BA are low adherence to therapy (43%), lack of elimination of triggers (29%), presence of comorbidities (15%), smoking (15%) and others [6]. Single nucleotide substitutions in the genome account for the impact of genetic polymorphism on the disease phenotype, and predetermine differences in the clinical manifestations of the disease, including symptom control.

A characteristic feature of molecular medicine as a science based on data on the molecular structure of the human genome is its focus on the correction of the pathological process in a particular person, taking into account his or her unique genetic features. Another peculiarity is preventive orientation, when information about genome obtained long before obvious manifestations of a disease can prevent its development. Genetic predisposition can manifest itself in interaction with environmental factors, that forms a pathological phenotype.

The most common method of studying the contribution of genetic mechanisms to the development of BA is to look for associations of the disease and its phenotypes with polymorphic markers of candidate genes.

Over the last decade, genetic studies have identified numerous candidate genes, which predispose to BA. However, the results reported in the literature are often contradictory, that confirms the need to further investigate the associations of BA with polymorphic markers of candidate genes.

Some polymorphic markers of candidate genes may contribute to the development of airway obstruction due to loss of lung elasticity, while others contribute to chronic inflammation leading to airway obstruction or poor response to medications such as β2-agonists or (iGCS) [7].

Genetic aspects of BA control in children continue to be studied. In particular, it is known that polymorphism of the β2-adrenergic receptor gene (ADRB2 gene) is associated with the therapeutic response of patients to bronchodilators - β2-agonists. Stimulation of β2-adrenergic receptors leads to bronchodilation and improvement of bronchial conduction, and also affects T-cell function, eosinophilic inflammation and causes a decrease in the secretion of proinflammatory mediators from mast cells [8].

The product of the HHIP gene (Hedgehog family interacting protein gene) is an evolutionarily conserved signaling protein, playing an important role in a wide range of processes. There is evidence of an association of the single-nucleotide polymorphism rs1828591 of the HHIP gene with predisposition to the development of bronchial obstruction [9]. The association of the single nucleotide polymorphism rs1512288 of the HHIP gene with reversibility of bronchial obstruction has been reported, while no such association has been found with bronchial hyperresponsiveness [10].

Among a significant number of BA candidate genes, cytokine-alarmin genes, which play a key role in all stages of allergic reactions, occupy a special place. In the direction of molecular genetic methods, the interleukin-33 (IL-33) cytokine-alarmin gene is promising for research. IL-33 is one of the central signaling molecules of immune reactions in BA [11]. The role of IL-33, has been confirmed in the pathogenesis of BA in children [12,13]. IL-33 levels have been found to be elevated in the sputum and bronchial biopsy specimens of asthmatic patients [14]. IL-33 is a tissue-derived cytokine, which induces and enhances eosinophilic inflammation and has emerged as a promising new target for the treatment of asthma and allergic diseases [15].

Binding of IL-33 to its receptor (ST2) increases the expression of several pro-inflammatory mediators (IL-5, IL-4 and IL-13), and thereby affects Th2-cell-mediated eosinophilic airway inflammation. Increased IL-33 was associated with the presence of the rs1342326 allele of the IL-33 gene. The IL-33 gene rs1342326 polymorphism was associated with a lower risk of asthma in children in Tunisian population and higher expression of IL-33 cytokine [16]. Single nucleotide polymorphism rs992969 of IL-33 gene, was reported to be associated with blood eosinophil levels, asthma and eosinophilic asthma. The polymorphism rs4008366 of IL-33 gene, showed a weak association with eosinophilic asthma [17].

A whole-genome sequencing in the Icelandic population revealed a rare variant of the IL-33 gene (NM_001199640:exon7:c.487-1G>C (rs146597587-C), allele frequency = 0.65%), which disrupts the canonical acceptor splice site upstream of the last coding exon of the gene [18]. This variant also occurs with low frequency in European population and is associated with lower eosinophil counts and reduced risk of asthma in Europeans (OR = 0.47; 95%). Heterozygotes have approximately 40% lower total IL-33 mRNA expression than non-carriers. This polymorphism results in the formation of a shortened form of IL-33 protein. The shortened variant does not form an IL-33R/ST2 complex and does not activate ST2-expressing cells. These data demonstrate that rs146597587-C is a loss-of-function cytokine mutation [18].

Despite a number of reported studies of HHIP, ADRB2 and IL-33 gene polymorphisms in BA, the significance of the association of polymorphisms of the above genes with the clinical course of BA in children has not yet been definitively determined. In connection with the above-mentioned, there is a need to improve the comprehensive assessment of the degree of BA control and to determine the influence of clinical, laboratory, functional and genetic characteristics on it.

### Aim of the study

To investigate the association of polymorphic variants of HHIP, ADRB2 and IL-33 genes with the phenotypes of clinical course of BA in children and the effectiveness of therapy of the disease

## Materials and Methods

A cohort single-center study of 90 bronchial asthma patients aged 5 to 17 years with an established diagnosis of bronchial asthma of varying degree of severity and control between November 2019 and March 2021 was conducted. The diagnosis of BA was established on the basis of current clinical guidelines. The study was conducted on the basis of the Regional Children’s Clinical Hospital of Perm and polyclinics of the Perm region within the framework of the RFBR research grant.

Inclusion criteria: children with diagnosed BA aged from 5 to 17 years;

Exclusion criteria: any acute respiratory infections at the time of examination, age less than 5 years (due to the impossibility of spirography in this age group).

All participants underwent a comprehensive examination of clinical condition and external respiratory function. Subsequently, a set of diagnostic procedures was implemented for them, including the study of genetic polymorphism of HHIP, ADRB2 and IL-33 genes to establish the relationship with clinical phenotypes, laboratory and instrumental examination parameters determining the course of bronchial asthma and the degree of disease control. All patients were collected anamnesis of the disease, including allergological anamnesis, general blood analysis, rhinocytogram, level of total and specific IgE, immunogram as indicated. Spirographic examination, pulse oximetry, chest X-ray examination, peak expiratory flow rate indices were performed. Based on the clinical parameters and respiratory function, the severity degree of exacerbation by clinical guidelines was identified.

The severity of BA exacerbations was determined according to the following clinical criteria - clinical symptoms, PSV, respiratory rate, pulse rate, frequency of emergency medication use, and nocturnal awakenings. The level of BA control was determined according to Asthma Control Test - ACT and C-ACT and the Composite Asthma Severity Index (CASI).

The material for the molecular genetic study was DNA isolated from dried capillary blood spots from 90 children. The allele and genotype frequencies of polymorphic loci of the following genes were studied: rs12551256-A and rs146597587-G of IL-33 gene in 70 children; rs12504628 of HHIP gene and ARG16GLY rs1042713 of ADRB2 gene in 90 BA patients with regard to the severity and control of the disease. The polymerase chain reaction (PCR) method was used to detect mutant alleles of genes. In children with severe BA, as well as in children with poorly controlled/uncontrolled asthma (n=26), sequencing of the entire coding sequence of the IL-33 gene located on the 9th chromosome in the 9p24.1 region was additionally performed (search for mutations in 9 exons).

The selection of cases was carried out by the method of continuous sampling. The methodology for determining the sample size was based on the use of a specialized formula, when the size of the general population is unknown:

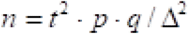

where n is the sample size, t is the coefficient depending on the confidence level chosen by the researcher, p is the proportion of respondents with the presence of the trait under study,

q = 1-p - the proportion of respondents who do not have the characteristic under study,

Δ - marginal sampling error.

Statistical processing of the results was carried out using Microsoft Excel 2010 statistical software packages.

The hypothesis of normality of distribution of the studied indicators was tested using the Shapiro-Wilk criterion. In order to extend the conclusions to the general population (95% reliability), some indicators were presented as M ±2m (% ±2m).

The two-sample Student’s t-test or its nonparametric analogue, the Mann-Whitney U-test was used to compare the dependent and independent groups of features characterizing the level of control and/or severity of the disease (depending on the type of distributions of the analyzed parameters). Pearson’s χ2 test was used to analyze the conjugation tables. The relationship between the variables was studied using correlation analysis. The significance level for the tested hypotheses was assumed to be 0.05.

The expected distribution of genotypes was estimated using the Hardy-Weinberg formula:

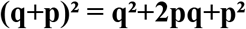

where q is the frequency of occurrence of recessive gene, p is the frequency of occurrence of dominant gene, q^²^ is the frequency of occurrence of genotype aa, p^²^ is the frequency of occurrence of genotype AA;

2pq is the frequency of occurrence of genotype Aa.

The χ2 test was used to assess whether the observed genotype distribution matched the expected one based on Hardy-Weinberg equilibrium.

For statistically significant parameters, relative risk (RR) was calculated using the formula:

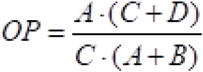

The confidence (95%) interval for the OR was calculated using the formulas:

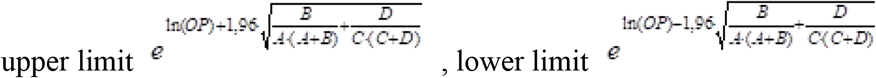

A, B, C, D in the formulas are the number of observations in the cells of the four-field conjugation table.

### Ethical review

This study was conducted in accordance with the principles of the World Medical Association Declaration of Helsinki. Informed consent was obtained in writing from the child’s legal representative in accordance with local laws and regulations before the child was included in the study. Conclusion of the local ethical committee at PSMU No. 5/20 dated 4 August 2020.

## Results and Discussion

All children of the study group were on pulmonologist’s dispensary registration with the diagnosis of bronchial asthma. The median age of the patients was 13 years [Q1-Q3: 9; 15 years]. A total of 100 children participated in the study, of whom 72 (72%) were boys and 28 (28%) were girls. In 62 patients BA had a mild course, 27 children were observed with moderate BA and 11 children with severe asthma. The largest number of patients in the study cohort (67 patients) had incomplete control of bronchial asthma, 7 children were diagnosed with no control and 26 patients had complete control.

When collecting anamnestic data, 87% of children were found to have hypoallergenic factors (presence of pets, carpets, flowering indoor plants, mould, passive smoking, gassing of the place of residence with exhaust fumes from cars or nearby industrial plants).

Outpatient medical records of children under dispensary supervision by a district paediatrician were analyzed to assess the comorbid background. Having studied the comorbidities of the examined children, allergic rhinitis - 80.0%, atopic dermatitis - 32.0% and pollinosis - 40.4% accounted for the highest percentage (Table 1). Most of the children had two or more comorbidities.

**Table 1:** Comorbidities in children with BA.

| Associated diseases | Number of children with established diagnosis, abs. |
| --- | --- |
| Allergic rhinitis | 80 |
| Atopic dermatitis | 32 |
| Pollinosis | 40 |
| Food allergy | 20 |
| Diseases of the gastrointestinal tract (chronic gastroduodenitis/ biliary dysfunction/chronic constipations) | 17 |
| Overweight/obesity | 15 |
| Congenital malformations of the tracheobronchial tree (TBT): extra bronchus/transposition of bronchus | 16 |
| Urticaria | 14 |

In the structure of complaints of the examined children, there dominated dyspnoea during physical activity (70.0 ± 9.0), dyspnoea outside in spring and summer (34.0 ± 9.3), respiratory infection (42.0 ± 9.7), labored breathing with wheezing (45.0 ± 9.8) and dry attack-like cough (48.0 ± 9.8).

The examination of external respiratory function using spirography revealed obstructive type disorders in 29 children, and 18% of children showed significant deterioration of parameters and their shift to the “red” zone, that once again confirms the hypothesis about the lack of disease control in BA patients on the background of baseline therapy.

### Characteristics of the studied genotypes

The distribution of genotypes in BA patients corresponded to the Hardy-Weinberg equilibrium, except for the genetic variant rs146597587 of IL-33 gene (G>C), where only carriers of one GG genotype were present (Table 2).

**Table 2:** Distribution of genotype frequencies of the studied polymorphisms (rs12504628 (T>C), rs1042713 (G>A), rs12551256, rs146597587 (G>C)) in the group of bronchial asthma patients.

| Gene/<br>Polymorphism | Genotype | N.O. | N.E. | $\chi^2$<br>df=1 | Allele<br>frequency | $h_{obs} \pm SE$<br>$h_{exp} \pm SE$ | D |
| --- | --- | --- | --- | --- | --- | --- | --- |
| *HHIP gene*<br>rs12504628<br>(T>C) | TT | 21 | 18,68 | 0,254<br>p=0,614 | T=0,455<br>C=0,544 | $h_{obs}=0,444 \pm 0,052$<br>$h_{exp}=0,101 \pm 0,012$ | –<br>0,586 |
|  | TC | 40 | 44,64 |  |  |  |  |
|  | CC | 29 | 26,68 |  |  |  |  |
|  | T | 82 | 45,56 |  |  |  |  |
|  | C | 98 | 54,44 |  |  |  |  |
| *ADRB2 gene*<br>rs1042713<br>(G>A) | GG | 40 | 39,34 | 0,004<br>p=0,952 | G=0,661<br>A=0,339 | $h_{obs}=0,433 \pm 0,052$<br>$h_{exp}=0,685 \pm 0,049$ | –<br>0,368 |
|  | GA | 39 | 40,33 |  |  |  |  |
|  | AA | 11 | 10,34 |  |  |  |  |
|  | G | 119 | 66,11 |  |  |  |  |
|  | A | 61 | 33,89 |  |  |  |  |
| *IL-33 gene*<br>rs12551256<br>(A>G) | GG | 24 | 22,56 | 0,095<br>p=0,758 | G=0,593<br>A=0,407 | $h_{obs}=0,438 \pm 0,062$<br>$h_{exp}=0,825 \pm 0,047$ | –<br>0,470 |
|  | GA | 28 | 30,88 |  |  |  |  |
|  | AA | 12 | 10,56 |  |  |  |  |
|  | G | 76 | 59,38 |  |  |  |  |
|  | A | 52 | 40,63 |  |  |  |  |
| *IL-33 gene*<br>rs146597587<br>(G>C) | GG | 40 | 100 | | G=1,0<br>C=0 | $h_{obs}=0$<br>$h_{exp}=0$ | 0 |
|  | CG | 0 | 0 |  |  |  |  |
|  | CC | 0 | 0 |  |  |  |  |
|  | G | 80 | 100 |  |  |  |  |
|  | C | 0 | 0 |  |  |  |  |

Note. N.O. - observed genotype abundance; N.E. - expected number of genotypes; $\chi^2$ criterion is used to assess whether the observed genotype distribution corresponds to the expected one based on Hardy-Weinberg equilibrium; df is the number of degrees of freedom; $h_{obs} \pm s.e.$ and $h_{exp} \pm s.e.$ - the observed and expected heterozygosity with error, respectively; D is the relative deviation of the observed heterozygosity from the expected one.

### Search for associations of genetic markers of HHIP, ADRB2 and IL-33 genes with the clinical course of bronchial asthma

Comparison of genetic markers in patients with severe BA (tBA) and non-severe BA (nBA) revealed a reduced risk of severe disease realization among those carrying the TT genotype (OR=0.221 (95% CI: 0.059-0.828; χ2=5.759; p=0.056)) and the T allele (OR=0.491 (95% CI: 0.190-1.269; χ2=4.270; p=0.039)) of the studied genetic variant rs12504628 (T>C) of the HHIP gene, the frequency of the CC genotype in severe BA was 64%, versus 28% in nonsevere BA, and the C allele 77% versus 52% (Table. 3).

**Table 3:** Case-control analysis of the studied genetic variants in severe and nonsevere BA.

| Gene/<br>Polymorphism | Genotypes/<br>alleles | BA<br>severe | | BA<br>nonsevere | | $\chi^2$ | p | OR |
| --- | --- | --- | --- | --- | --- | --- | --- | --- |
|  |  | N | % | N | % |  |  |  |
| *HHIP gene*<br>rs12504628<br>(T>C)<br>1-T/C, 2-T/T,<br>3-C/C | TT | 1 | 9 | 20 | **25** | 5,759 | 0,056 | **0,221**<br>(0,059<OR<0,828) |
|  | TC | 3 | 27 | 37 | 47 |  |  |  |
|  | CC | 7 | 64 | 22 | 28 |  |  |  |
|  | T | 5 | **23** | 77 | **48** | **4,270** | **0,039** | **0,309**<br>(0,109<OR<0,880) |
|  | C | 17 | 77 | 81 | 52 |  |  |  |
| *ADRB2 gene*<br>rs1042713<br>(G>A)<br>1-G/A, 2-G/G,<br>3-A/A | GG | 5 | 45 | 35 | 44 | 1,898 | 0,387 | 1,048<br>(0,295<OR<3,720) |
|  | GA | 6 | 55 | 33 | 42 |  |  |  |
|  | AA | 0 | 0 | 11 | 14 |  |  |  |
|  | G | 16 | 73 | 103 | 65 | 0,211 | 0,646 | 1,424<br>(0,527<OR<3,847) |
|  | A | 6 | 27 | 55 | 35 |  |  |  |
| *IL-33 gene*<br>rs12551256<br>(A>G)<br>1-A/G, 2-A/A,<br>3-G/G | GG | 3 | 11 | 21 | 14 | 4,345 | 0,114 | 0,470<br>(0,190<OR<1,166) |
|  | GA | c | 45 | 28 | 40 |  |  |  |
|  | AA | c | 44 | 12 | 46 |  |  |  |
|  | G | 6 | 33 | 70 | 34 | 0,027 | 0,870 | 0,950<br>(0,646<OR<1,396) |
|  | A | 0 | 67 | 52 | 66 |  |  |  |
| *IL-33 gene*<br>rs146597587<br>(G>C)<br>1-G/C, 2-G/G,<br>3-C/C | GG | 8 | 11 | 22 | 14 | 4,345 | 0,114 | 0,470<br>(0,190<OR<1,166) |
|  | GC | 0 | 45 | 0 | 40 |  |  |  |
|  | CC | 0 | 44 | 0 | 46 |  |  |  |
|  | G | 16 | 33 | 44 | 34 | 0,027 | 0,870 | 0,950<br>(0,646<OR<1,396) |
|  | C | 0 | 67 | 0 | 66 |  |  |  |

Note: N - absolute number of observed genotypes, p is given for $\chi^2$ test, c - sequencing.

Comparison of genetic markers in patients with a combination of atopic dermatitis (AtD) and bronchial asthma (BA+AtD) and BA without AtD (BA without AtD) revealed an increased risk of combining asthma and dermatitis among individuals carrying the TT genotype (OR=2.875 (95% CI: 1.130-7.316; χ2=5.751; p=0.056)) of genetic variant rs12504628 (T>C) of the HHIP gene.

Analysis of genetic markers in patients with a combination of congenital malformations (CM) of the tracheobronchial tree with asthma (BA+CM) and bronchial asthma without malformations (BA without CM) revealed associations with the combination of asthma and CM with the AA genotype (OR=0.182 (95% CI: 0.051-0.646; χ2=8.567; p=0.014)) genetic variant rs1042713 (G>A) of the ADRB2 gene, which has a protective value against CM, it was shown that AA genotype carriers had bronchial tree CM in 40% of cases, compared to 11% of AA genotype carriers without CM.

Analysis of genetic markers in patients with BA and aggravated allergy history among 1st degree relatives (BA+ hereditary history) and bronchial asthma without aggravated hereditary allergy history (BA without hereditary history) revealed that AA genotype carriers (OR=0.112 (95% CI: 0.013-0.932; χ2=5.554; p=0.062)) and carriers of the A allele (OR=0.453 (95% CI: 0.213-0.964; χ2=3.537; p=0.059)) genetic variant rs1042713 (G>A) of the ADRB2 gene, have a lower incidence of an aggravated allergy history among 1st degree relatives (4% vs 26% for genotype AA and 26: vs 44% for allele A).

Analysis of genetic markers in patients with BA and drug allergy (BA+DA) and bronchial asthma without drug allergy (BA without DA) revealed that carriers of the CC+TS genotype were more likely to have a combination of asthma and drug allergy (OR=2.917 (95% CI: 1.009-8.427; χ2=4.984; p=0.083)) genetic variant rs12504628 (T>C) of the HHIP gene, a protective role was shown for the T allele in relation to the realization of drug allergy (31% vs 52%) (OR=0.416 (95% CI: 0.190-0.909; χ2=4.204; p=0.040)).

Analysis of genetic markers in patients with uncontrolled BA and bronchial asthma with partial and complete control of disease symptoms showed no associations with the genetic variants studied.

Carrying the TT genotype of genetic variant rs12504628 (T>C) of the HHIP gene reduces the risk of severe BA, but increases the risk of concomitant atopic dermatitis by 2.8 times. Carriage of CC+CT genotypes increases the risk of drug allergy on the background of BA by 2.9 times (Table 4).

**Table 4:** Associations of bronchial asthma with genetic variants studied.

| Gene/<br>Polymorphism | Comparison<br>genotype/allele | OR (95%<br>CI) | Clinical associations |
| --- | --- | --- | --- |
| *HHIP* gene<br>rs12504628<br>(T>C) | TT vs CC+TC | 0,221<br>(0,059-0,828) | Reduced risk of severe BA realization for TT genotype carriers |
|  | C vs T | 0,309<br>(0,109-0,880) | Reduced risk of severe BA realization for carriers of the T allele |
|  | TT vs CC+TC | 2,875<br>(1,130-7,316) | Increased risk of combining BA and atopic dermatitis for carriers of TT genotype |
|  | CC+TC vs TT | 2,917<br>(1,009-8,427) | Increased risk of combining BA and drug allergy for CC+TC genotype carriers |
|  | C vs T | 0,416<br>(0,190-0,909) | Reduced risk of drug allergy against the background of BA for carriers of the T allele |
| *ADRB2* gene<br>rs1042713<br>(G>A) | AA vs GA+GG | 0,182<br>(0,051-0,646) | Reduced risk of bronchial tree allergy against the background of BA for AA genotype carriers. |
|  | AA vs GA+GG | 0,112<br>(0,013-0,932) | Decrease in the frequency of aggravated allergic anamnesis against the background of BA for AA genotype carriers. |
|  | A vs G | 0,453<br>(0,213-0,964) | Decrease in the frequency of aggravated allergic anamnesis against the background of BA for carriers of the AA allele. |

Carrying the AA genotype of the ADRB2 gene is associated with a decreased risk of allergic history among 1st degree relatives and a decreased risk of congenital malformations of the tracheobronchial tree against the background of BA (Table 4).

Sequencing and exome analysis of the IL-33 gene showed a statistically significant positive association between the frequency of lesions in exons 4 (r=0.417; p=0.034) and 6 (r=0.593; p=0.001) on the one hand and the severity of BA on the other. Nucleotide substitutions in these exons were found to be more frequently associated with the severe course of bronchial asthma.

The study [14] shows that the homozygous variant on Arg16 of the ADRB2 gene is associated with an increased risk of developing a more severe form of bronchial asthma, as well as its nocturnal form compared to homozygotes on the Gly16 allele. An analysis of 28 published studies on the association of beta2-adrenoreceptor gene polymorphisms with BA phenotypes confirmed an association between the Gly16 polymorphism and nocturnal asthma, but no association was found between the Arg16Gly variant and bronchial hyperresponsiveness [20, 21]. In our study, we were able to confirm that the AA genotype of the ADRB2 gene was associated with a decreased risk of aggravated allergoanamnesis among 1st degree relatives and a decreased risk of realization of congenital malformations of the tracheobronchial tree against the background of BA.

It should be noted that children with the indicated AA genotype of the ADRB2 gene in our sample did not receive long-acting b2-agonists as monotherapies of baseline therapy and did not use monotherapy with short-acting b2-agonists as emergency drugs during the last 6 months. Consequently, it is not possible to assess the risk of disease exacerbation in Arg16 genotype carriers who resort to the use of b2-agonists.

The HHIP gene has now been found to affect both small and large airways [22]. The presence of allele A of the rs13118928 polymorphism of the HHIP gene may be associated with the emphysema-hyperinflation phenotype in patients with chronic obstructive pulmonary disease [23]. It is known that the state of external respiratory function is the most important criterion of bronchial asthma severity. We found that carrying the TT genotype of the genetic variant rs12504628 (T>C) of the HHIP gene reduces the risk of severe BA, but increases the risk of atopic dermatitis combined with BA by 2.8 times. The presence of CC+CT genotypes increases the risk of drug allergy against the background of BA by 2.9 times.

On the one hand, we did not detect any association of genetic variants of the IL-33 gene rs12551256 polymorphism with the clinical course of BA. When analyzing the genotypes of the rs146597587 (G>C) polymorphism of the IL-33 gene, all children were carriers of the same genotype. On the other hand, nucleotide substitutions in exons 4 and 6 of the IL-33 gene were found to be associated with a severe course of bronchial asthma. This justifies the expediency of further study of IL-33 gene exon polymorphism and its associations with the clinical course of bronchial asthma in children with an increased patients’ sample size.

## Study limitations

The association of HHIP, ADRB2 and IL-33 gene polymorphisms in children cannot be extrapolated to the entire population of Russian children due to the small size of the study sample. It is possible that when the sample size is increased, the distributions of genotypes and alleles of these genes will differ from those presented in this article. We did not perform multivariate analysis with correction for the detected gene associations, taking into account the carriage of polymorphic variants of other genes and environmental factors, that may affect the results of the evaluation of the effect of the studied genes.

## Conclusion

The study of HHIP, ADRB2 and IL-33 gene polymorphisms in BA children with different disease phenotypes revealed an association between gene polymorphisms and disease severity, as well as with comorbidities.

It was shown that TT genotype of genetic variant rs12504628 (T>C) of HHIP gene reduces the risk of severe BA, but increases the risk of atopic dermatitis combined with BA by 2.8 times. The CC+ST genotype of the HHIP gene increases the risk of drug allergy against the background of BA by 2.9 times.

The AA genotype of the ADRB2 gene is associated with the absence of an aggravated allergoanamnesis among 1st degree relatives and a decreased risk of congenital malformations of the tracheobronchial tree against the background of BA. Polymorphic variants in exons 4 and 6 of the IL-33 gene are more often combined with moderate and severe asthma, and nucleotide substitutions in exons 4 and 6 are associated with a severe course of BA. This suggests that exons 4 and 6 should be paid attention to in clinical practice to predict the course of the disease and timely correction of baseline therapy.

Thus, this study has established associations of polymorphic variants of HHIP, ADRB2 and IL-33 genes with clinical manifestations of bronchial asthma in children, which can be taken into account in the personalized monitoring of these patients and can help to achieve complete control of the disease.

This research was supported by a joint grant from the Israel Ministry of Science and Technology (MOST, 3-16500), the Russian Centre for Scientific Information (RFBR) (Joint Research Project 19-515-06001)

## Data Availability

All data produced in the present study are available upon reasonable request to the authors

